# Cohort Profile: The Danish National Cohort Study of Effectiveness and Safety of SARS-CoV-2 vaccines (ENFORCE)

**DOI:** 10.1101/2022.10.09.22280886

**Authors:** Nina Breinholt Stærke, Joanne Reekie, Isik S. Johansen, Henrik Nielsen, Thomas Benfield, Lothar Wiese, Ole Schmeltz Søgaard, Martin Tolstrup, Kasper Karmark Iversen, Britta Tarp, Fredrikke Dam Larsen, Lykke Larsen, Susan Olaf Lindvig, Inge Kristine Holden, Mette Brouw Iversen, Lene Surland Knudsen, Kamille Fogh, Marie Louise Jakobsen, Anna Traytel, Lars Østergaard, Jens Lundgren, the ENFORCE Study Group

## Abstract

**Purpose:** The ENFORCE cohort is a national Danish prospective cohort of adults who received a Severe acute respiratory syndrome coronavirus 2 (SARS-CoV-2) vaccine as part of the Danish National SARS-CoV-2 vaccination program. It was designed to investigate the long-term effectiveness, safety and durability of SARS-CoV-2 vaccines used in Denmark.

**Participants:** A total of 6943 adults scheduled to receive a SARS-CoV-2 vaccine in the Danish COVID-19 Vaccination Program were enrolled in the study prior to their first vaccination. Participants will be followed for a total of two years with five predetermined follow-up visits and additional visits in relation to any booster vaccination. Serology measurements are performed after each study visit. T-cell immunity is evaluated at each study visit for a subgroup of 699 participants. Safety information is collected from participants at visits following each vaccination. Data on hospital admissions, diagnoses, deaths and SARS-CoV-2 polymerase chain reaction (PCR) results are collected from national registries throughout the study period. The median age of participants was 64 years (IQR 53-75), 56.6% were females and 23% were individuals with an increased risk of a serious course of COVID-19. A total of 340 (4.9%) participants tested positive for SARS-CoV-2 spike IgG at baseline.

**Findings to date:** Results have been published on risk factors for humoral hyporesponsiveness and non-durable response to SARS-CoV-2 vaccination, the risk of breakthrough infections at different levels of SARS-CoV-2 spike IgG by viral variant, and on the antibody neutralizing capacity against different SARS-CoV-2 variants following primary and booster vaccinations.

**Future plans:** The ENFORCE cohort will continuously generate studies investigating immunological response, effectiveness, safety and durability of the SARS-CoV-2 vaccines.

**Registration:** clinicaltrials.gov identifier: NCT04760132.

**Strengths and limitations:**

- The ENFORCE study combines repeated detailed SARS-CoV-2 specific immunological measurements prior to, and throughout the course of SARS-CoV-2 vaccination, with register-based follow-up of safety data and microbiological test results.
- The ENFORCE cohort includes a large proportion of elderly participants and participants with concomitant diseases.
- The three vaccine groups display a high degree of variation in demographic factors and distribution across risk groups, due to the prioritization of specific vaccines to risk groups during the primary roll out of the SARS-CoV-2 vaccination program.

## Introduction

Upon the emergence of SARS-CoV-2, the pathogen causing COVID-19, the development of effective vaccines quickly became of highest priority for decision makers, researchers, and pharmaceutical companies throughout the world. Subsequently, within one year of the emergence of the virus, the first reports from clinical phase 1-2 trials of SARS-CoV-2 vaccines in humans were published ^1-5^.

The first SARS-CoV-2 vaccine approved for emergency use in Denmark was the Pfizer-BioNTech mRNA vaccine BNT162n2 (Pfizer Inc., New York, United States of America; BioNTech SE, Mainz, Germany) ^6^. It was used from the initiation of the Danish COVID-19 vaccination program on 27th of December 2020. In January 2021 both the Moderna mRNA-1273 vaccine (Moderna Inc., Massachusetts, United States of America) ^7^ and the AstraZeneca adenovirus-vectored ChAd0×1 nCoV-19 vaccine (AstraZeneca, Cambridge, United Kingdom) ^8^ were approved for emergency use. The adenoviral vectored vaccine Ad26.COV2.S (Johnson&Johnson, New Jersey, United States of America)^9^ was used in a supplementary program alongside the Danish National COVID-19 vaccination program and only a small group of individuals received this vaccine in Denmark.

As of September 2022, more than 600 million cases and 6 million deaths due to COVID-19 have been reported to the WHO. Globally, over 12 billion vaccine doses against SARS-CoV-2 have been administered, however especially on the African continent, large parts of the population are still lacking their first SARS-CoV-2 vaccination^10^. Conversely, in Denmark and similar countries, the 4^th^ vaccination dose is being planned for selected parts of the population^11^. Additionally, new Omicron specific and bivalent vaccines are being developed and introduced in vaccination programs replacing the initial vaccines^12,13^.

The efficacy and safety of the SARS-CoV-2 vaccines were described in defined study populations and smaller cohorts as the vaccines were introduced ^6-8,14-20^, however, there is an ongoing need for knowledge on the long-term effectiveness and safety of the vaccines, the durability and magnitude of vaccine induced immune response, protection against new emerging variants and the response to booster doses particularly in risk groups.

### Cohort Description

ENFORCE was designed as an open-labelled, non-randomized, parallel group, phase IV study enrolling adults residing in Denmark before their first SARS-CoV-2 vaccination offered through the Danish COVID-19 vaccination program. The inclusion period ran from the 13^th^ of February 2021 to the 5^th^ August 2021.

The study was planned by the ENFORCE Consortium and is being carried out as a collaborative effort with seven study sites in the cities Aalborg, Silkeborg, Aarhus, Odense, Roskilde, Hvidovre and Herlev covering all 5 Danish regions.

#### Study population

Participants had to be eligible for SARS-CoV-2 vaccination through the Danish COVID-19 Vaccination Program, willing and able to provide informed consent and willing and able to comply with trial procedures. Potential participants would be excluded if they were below 18 years of age, belonged to a subgroup for which the SARS-CoV-2 vaccines were contra-indicated or had received a previous SARS-CoV-2 vaccination.

Potential study participants were invited to join the ENFORCE study through a number of channels selected to ensure a high proportion of persons at increased risk of a serious course of SARS-CoV-2 infection, and health care workers (HCW) at risk of SARS-CoV-2 exposure. A letter of invitation was sent to selected risk groups such as cancer patients in active therapy, organ transplant recipients, patients with immunodeficiencies, patients in hemodialysis treatment and patients with certain rheumatologic and pulmonary diseases. HCW were invited to participate via hospital information channels. The general population was invited to participate through a letter of invitation sent via the vaccination centers to adults with a scheduled appointment for vaccination in any of the study site cities.

Enrollment visits were performed by ENFORCE study personnel, either at vaccination centers located near study sites, or at hospital clinics. Participants could be enrolled from 14 days up to 30 minutes before their first dose of a SARS-CoV-2 vaccine.

#### Data collection

At enrollment, baseline information was collected on age, sex, focused medical history, concomitant medication, vaccination priority group (defined by the Danish COVID-19 Vaccination Program ^21^), scheduled vaccination date and vaccine manufacturer (Pfizer-BioNTech, Moderna, AstraZeneca or Johnson&Johnson). Participants also provided blood samples for baseline SARS-CoV-2 serology. Persons at increased risk of a serious course of SARS-CoV-2 infection were classified as such, on the basis that they were referred to vaccination by a medical specialist tending to their disease. The group included cancer patients in active treatment, patients with immunodeficiencies (acquired or inherent), organ transplant recipients, hemodialysis patients, and some patients with hematological, pulmonary or rheumatological diseases. Individuals that were vaccinated because of an occupation as HCW were registered as such. Vaccine type was confirmed by cross referencing each individual’s unique civil registration (CPR) number with data from the Danish Vaccination Register (Statens Serum Institut, Copenhagen, Denmark).

Safety data is collected using a questionnaire (either online or physical) on solicited local and systemic reactions filled out by the participants at one and two weeks following each vaccination. Additionally, information regarding adverse events is collected until the visits following each vaccination. Data on specific concomitant diseases, newly diagnosed medical conditions, hospitalizations and death are collected from the Danish National Patient Register (Danish Health Data Authority, Copenhagen, Denmark).

Data on any SARS-CoV-2 PCR-tests or SARS-CoV-2 antibody measurements were extracted from the surveillance system Key Infectious Diseases System (KIDS, Statens Serum Institut, Copenhagen, Denmark), whereas viral variant information is obtained through the Danish National Microbiology Database (MiBa, Statens Serum Institut, Copenhagen, Denmark).

#### Follow-up

Study participants are followed with 5 predetermined clinic visits in addition to the baseline visit. The second study visit is scheduled at 0-5 days prior to the participants second SARS-CoV-2 vaccination, while the following visits are at 3, 6, 12 and 24 months after the first vaccination. Furthermore, study visits are scheduled 0-14 days before and one month after any SARS-CoV-2 booster vaccination. An outline of the study visit schedule is shown in figure 1.

**Figure 1:**
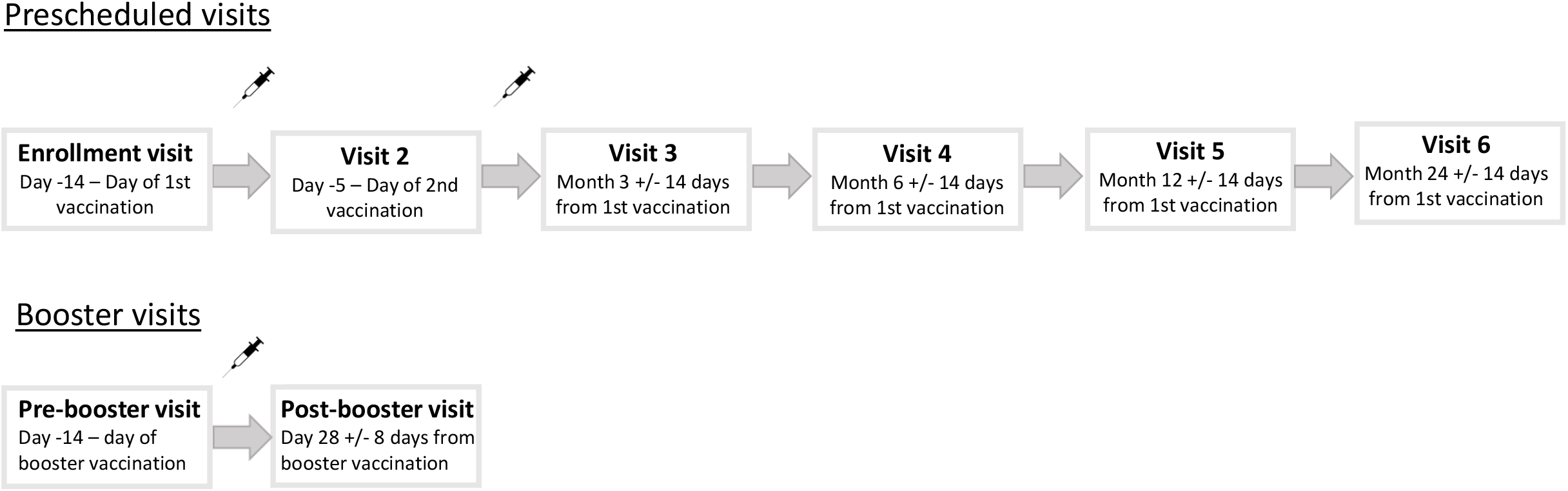
Schematic overview of the study visit schedule. Syringe icons signify SARS-CoV-2 vaccinations.

#### Serology measurements

At each study visit serum and plasma samples are collected from all participants for measuring specific SARS-CoV-2 serology. SARS-CoV-2 spike IgG is measured on serum samples utilizing a Wantai ELISA based assay (Beijing Wantai Biological Pharmacy Enterprise Co., Ltd., Beijing, China) performed at Statens Serum Institut, Copenhagen, Denmark. Additionally, a Multiantigen serology Assay (Meso Scale Diagnostics LLC, Maryland, United States of America) quantifying spike receptor binding domain, full spike and nucleocapsid directed IgG and ACE-2 competition is performed on plasma samples at the Research Laboratory at the Department of Infectious Diseases, Aarhus University Hospital, Aarhus, Denmark. Additional plasma and serum samples for future research analyses are collected and stored at the Danish National Biobank at Statens Serum Institute.

#### Cellular immune response

A group consisting of 699 participants evenly distributed across vaccine and age groups were enrolled in a sub-study evaluating SARS-CoV-2 spike specific T-cell immunity using peripheral blood mononuclear cells. For this subgroup additional blood samples, including RNA stabilizing PAXgene tubes, are collected at each visit. Analyses of T-cell immunity are performed at the Research Laboratory at the Department of Infectious Diseases, Aarhus University Hospital, Aarhus, Denmark.

#### Vaccine groups

The first participant included in this study received the AstraZeneca adenovirus vectored vaccine on the 13^th^ of February 2021. However, the use of the AstraZeneca vaccine in the Danish COVID-19 vaccination program was stopped on the 11^th^ of March 2021, and therefore only 474 participants received this vaccine. A small group of 25 participants received the adenovirus vectored vaccine manufactured by Johnson and Johnson. In total 499 participants received an adenovirus vectored vaccine at their first vaccination and are included in the study as the adenovirus vector/mRNA group.

Inclusion in the Pfizer-BioNTech group started on 16^th^ of February 2021. This group was temporarily paused from mid-April to mid-May 2021 when the initial target of 2500 participants was reached, but was subsequently resumed in order to enroll more participants in younger age groups and concluded at 3824 participants. Inclusion in the Moderna group started on the 24^th^ of February 2021, but was slow-paced in the beginning due to limited vaccine deliveries, but finalized at 2620 participants. Enrollment progression during the inclusion period for each vaccine group is shown in figure 2.

**Figure 2:**
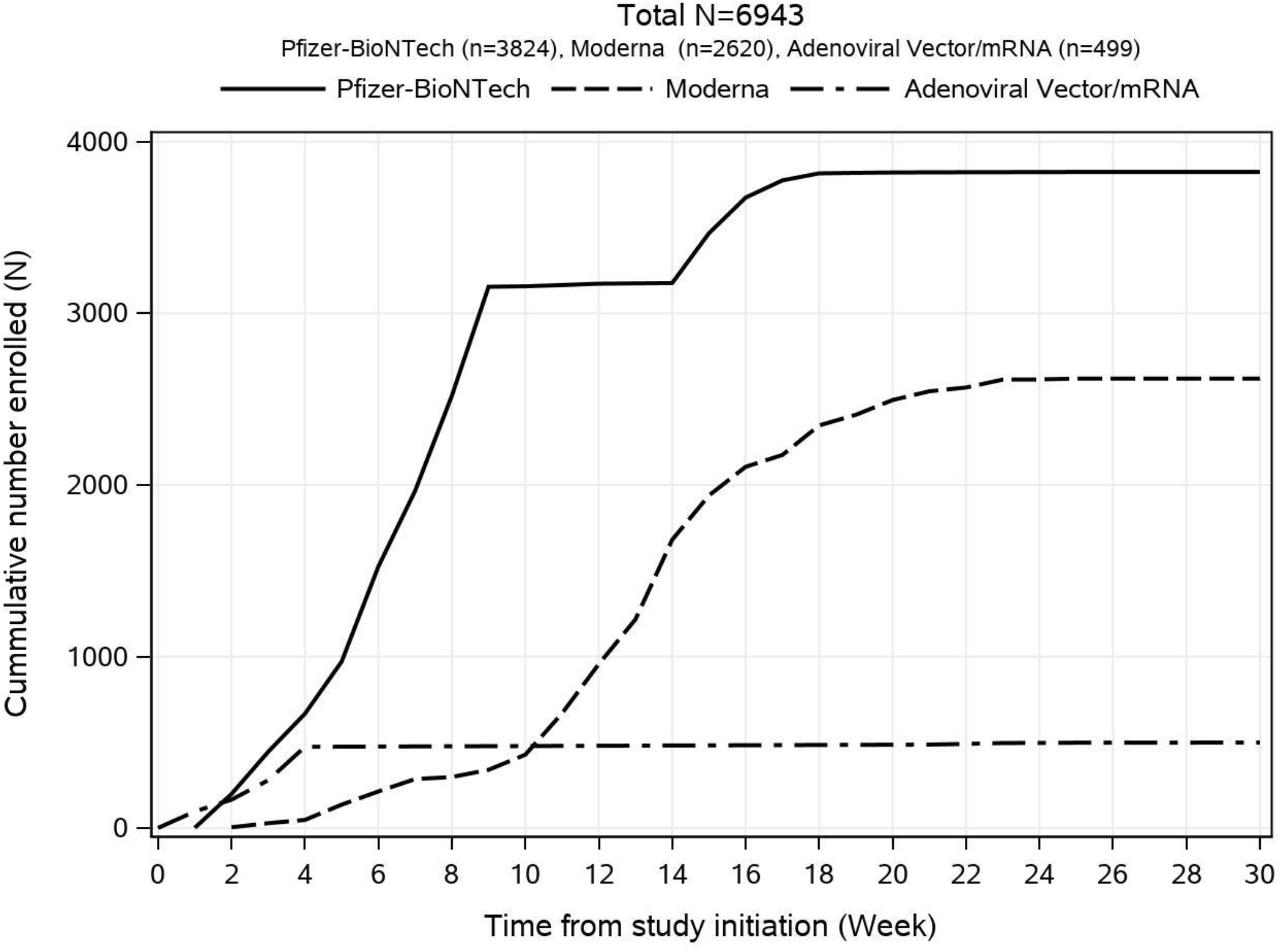
Cumulative number of participants enrolled over the inclusion period shown by vaccine type.

#### Demographics

A total of 6972 participants were enrolled in the study and 6943 participants (99.6%) are included in the analysis. The most common reason for exclusion was withdrawal of consent (n=23) and receiving a non-standard vaccine regimen (n=4). Reasons for exclusion are shown in figure 3.

**Figure 3:**
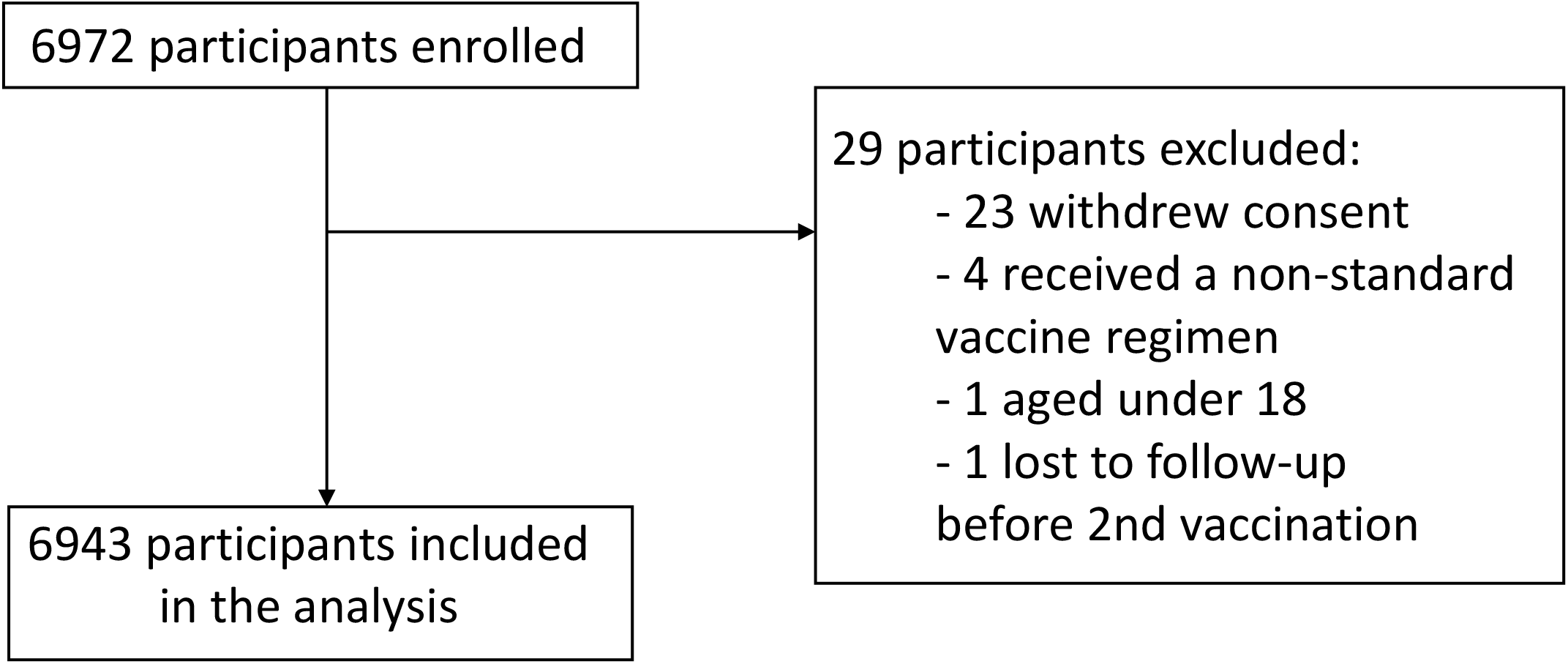
Flowchart of reasons for exclusion from the study.

Overall, the median age was 64 years (IQR 53-75 years), 3929 (56.6%) participants were females, 590 (8.5%) were HCW, and 1599 (23%) were classified as having an increased risk of a serious course of COVID-19 due to concomitant diseases. Overall, 3.4% (n=239) of the cohort had a positive SARS-CoV-2 PCR test prior to enrollment in the study. Participants in the adenoviral vector/mRNA group (N=499) were generally younger with a median age of 45 years (IQR 31-55 years) and 411 (82.4%) were female, while 453 (90.8%) were HCW. A higher proportion in the adenoviral vector/mRNA group (n=47, 9.4%) had a positive SARS-CoV-2 PCR prior to baseline compared with the other vaccine groups. The Pfizer-BioNTech group (N=3824) had a larger proportion of participants with an increased risk of a serious course of infection with SARS-CoV-2 (n=1437, 37.6%) and the participants were generally older, with a median age of 71 years (IQR 55-78 years), relative to the other vaccine groups. The demographic distribution overall and within the vaccine groups is summarized in table 1.

**Table 1.**
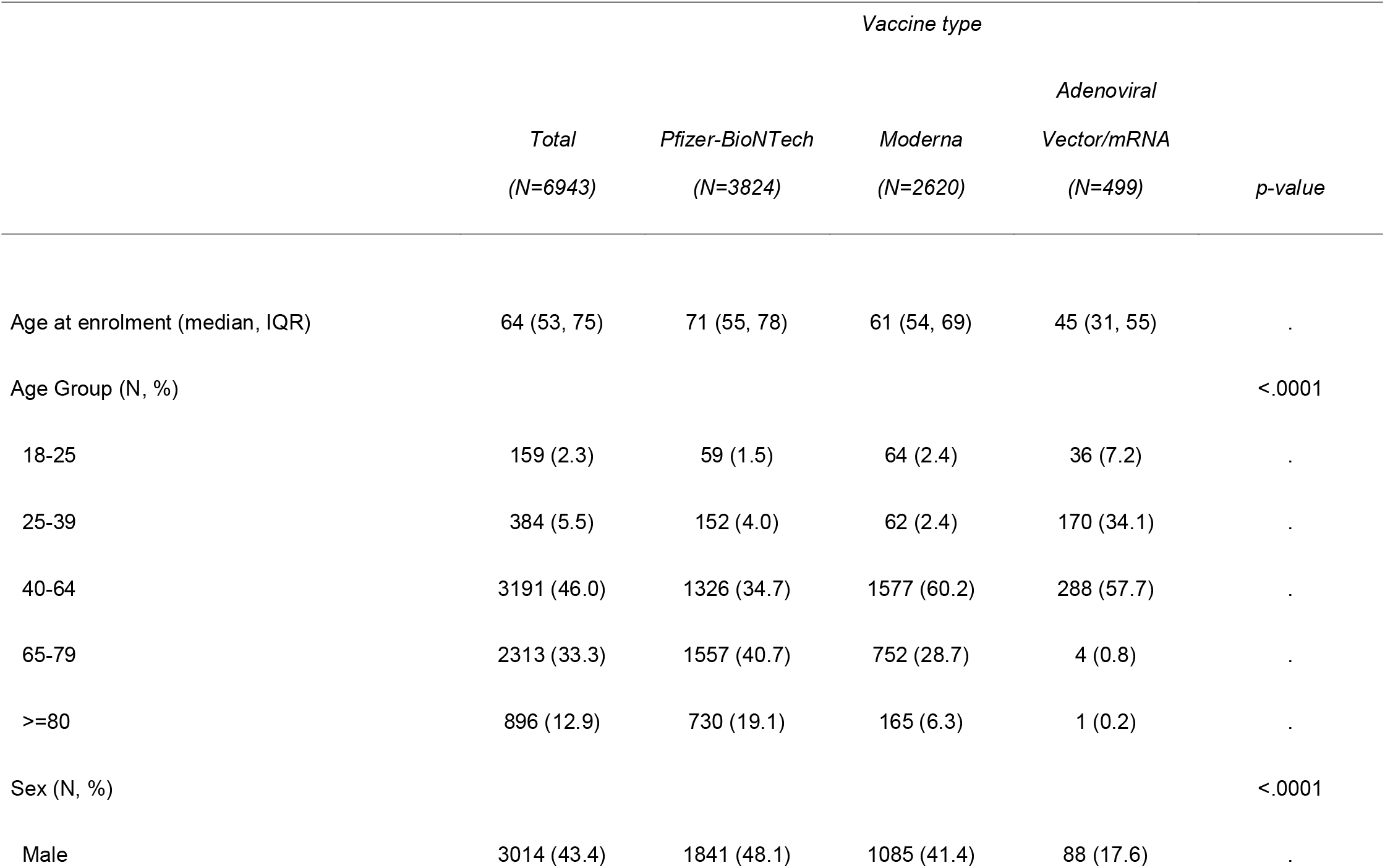
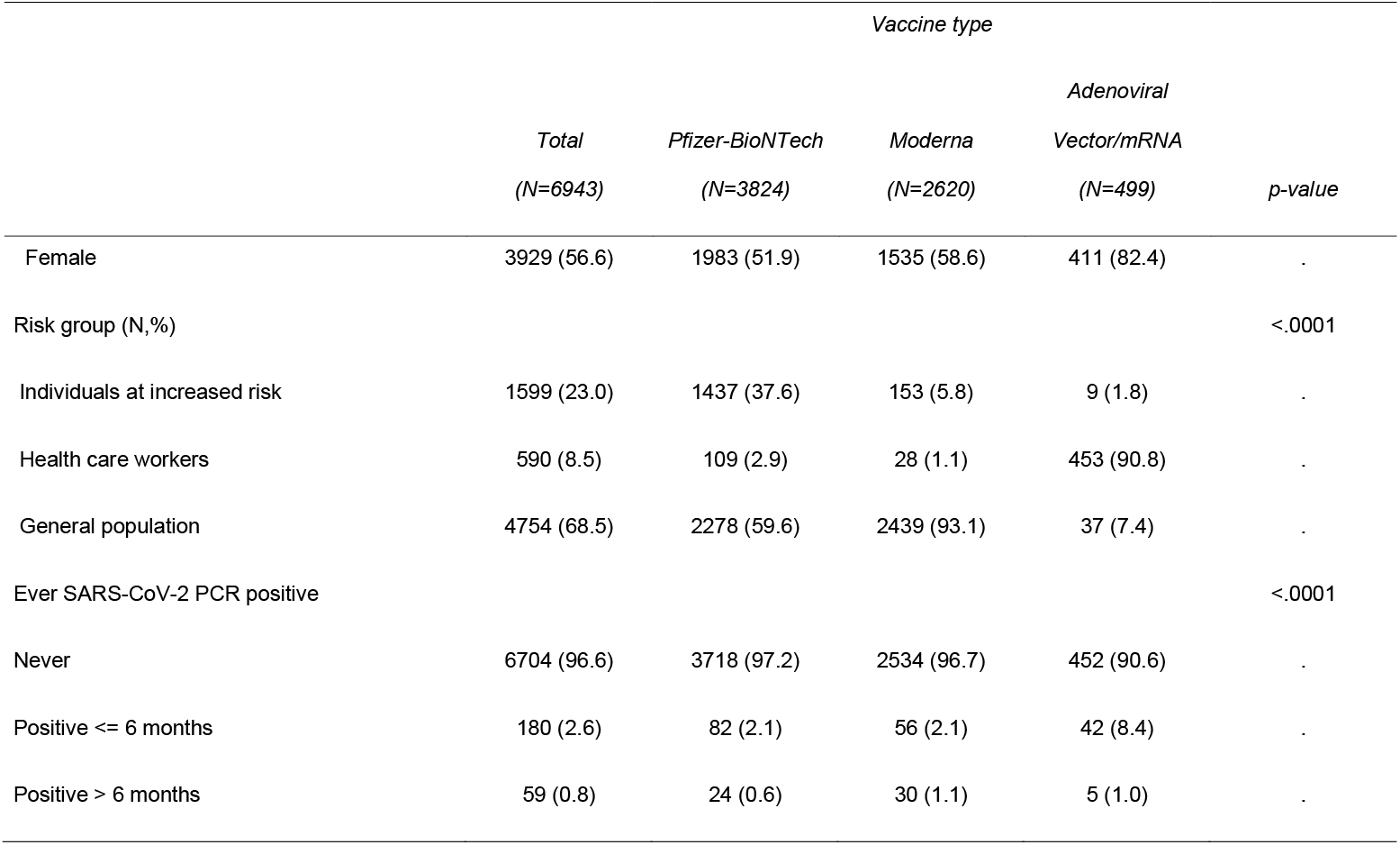
Participant demographics at study enrolment by vaccine type. Participant characteristics at baseline are presented overall and compared across vaccine groups, using either the Chi-squared test for categorical variables or Kruskal Wallis test for continuous variables.

#### Baseline Wantai serology results

A total of 6889 participants (99.2% of the analyzed population) had a conclusive result of the baseline Wantai analysis measuring SARS-CoV-2 spike IgG. Of those 340 participants (4.9%) had a positive result, indicating previous infection with SARS-CoV-2. The proportion of positive participants were 17/157 (10.8%) among the youngest participants aged 18-25 years, while it was 26/893 (2.9%) among those over the age of 80 years. Serology results are summarized in figure 4.

**Figure 4:**
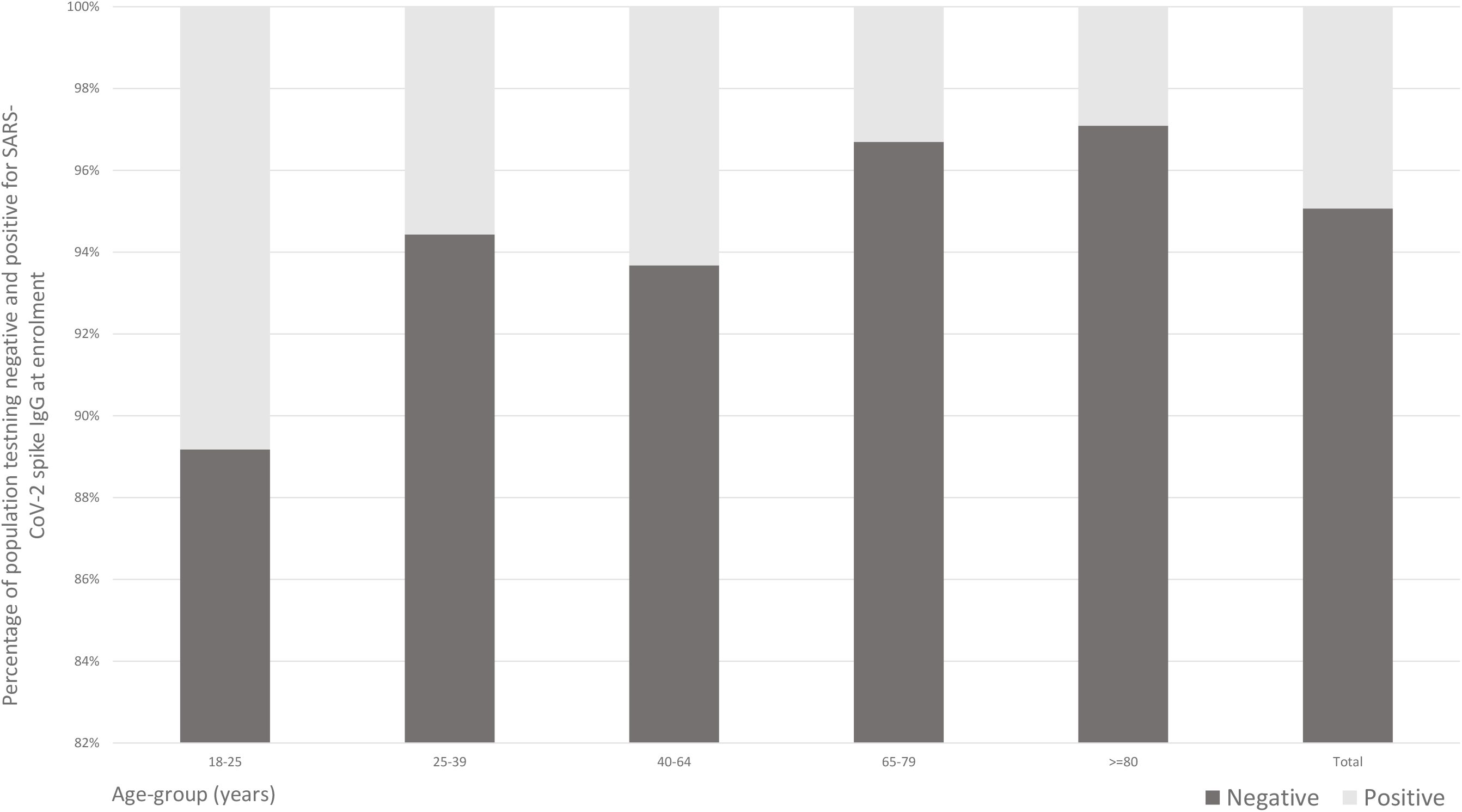
Percentage of population testing positive and negative for SARS-CoV-2 IgG by means of the Wantai analysis at enrolment, shown for age groups and overall.

#### Participant and public involvement

Public reports on the preliminary study results are shared on the ENFORCE website and a public seminar was held, where study results were presented. Participants in ENFORCE receive regular updates on the study progress and results, including results of their own antibody measurements.

### Findings to date

We assessed levels of SARS-CoV-2 spike IgG and Spike-ACE2-receptor-blocking antibodies at visits up to six months after the participants’ first SARS-CoV-2 vaccination and identified risk factors for humoral hyporesponsiveness and lack of durability of the vaccine response. We found that male sex, level of co-morbidity and vaccine type were all associated with hyporesponsiveness and non-durability of both antibodies assessed^22^. After the emergence of the Omicron variant, we conducted a study comparing the risk of breakthrough infection at different levels of SARS-CoV-2 spike IgG stratified by viral variant. We found an inverse association between increasing level of spike IgG and risk of breakthrough infection for the Delta variant, while there where no association between antibody levels and risk of breakthrough infection with the Omicron variant^23^. Recently, a study was published comparing the vaccine-induced antibody response and neutralizing capacity towards a range of SARS-CoV-2 variants after primary vaccination and first booster dose. Here it was found that the neutralizing capacity was reduced for each of the emergent variants tested as compared to wild type, and most pronounced for the omicron variant. However, it was also seen that neutralizing capacity increased significantly after the admission of a booster dose^24^.

### Strengths and limitations

The primary strength of the ENFORCE study is that it combines repeated measurements of SARS-CoV-2 specific humoral and cellular immunological response throughout the course of SARS-CoV-2 vaccination, with register-based follow-up of safety data and microbiological test results. This design allows the ENFORCE cohort to contribute with significant knowledge of the interplay between humoral and cellular immune response to SARS-CoV-2 vaccination and new emerging SARS-CoV-2 variants, hybrid immunity and long-term safety of SARS-CoV-2 vaccination.

The ENFORCE cohort has high proportions of participants over the age of 65 years and of participants with significant comorbidities. These groups are of particular interest in the context of vaccine effectiveness and durability as they have an increased risk of developing a serious course of COVID-19, and vaccine programs are often designed to protect these individuals.

The three vaccine groups display a large degree of variation in demographic factors and distribution across risk groups, which may be explained by the availability and prioritizing of specific vaccines to specific groups at different time points. The AstraZeneca vaccine was specifically prioritized for HCW before it was removed from the vaccination program, and therefore the adenovirus vectored/mRNA group includes higher proportions of HCW, females and younger age groups. The Pfizer-BioNTech vaccine was used to a large extent for the elderly population and individuals at increased risk of a serious course of disease, as it was the vaccine mainly available in the first months of the vaccination program. The between-group differences complicates the comparison of vaccine groups. A randomized design would have eliminated this limitation, but due to vaccine shortage in the early stage of the vaccination program and vaccine prioritization for risk groups it was unfeasible to randomize participants to vaccine groups.

The use of registry data comprises a risk of misclassification due to incorrect registration. However, the large size of the ENFORCE cohort reduces the risk of misclassification affecting results significantly.

### Future plans

Studies are planned or ongoing investigating the T-cell immune response of the cohort at different timepoints relative to primary vaccination and boosters, the significance of hybrid immunity, morbidity and mortality compared with those of a historic control group, and the correlation of local and systemic reactions to vaccination with antibody response. Data collection will proceed until July 2023.

### Collaboration

The ENFORCE study encourages scientific collaboration. While the study is ongoing data may be made available to scientists upon approval of an application sent to the ENFORCE Scientific Steering Committee and further approval by relevant authorities. Applications for data must be sent to. Detailed information about data access may be found here: https://chip.dk/Research/Studies/ENFORCE/Study-Governance.

## Supporting information

The ENFORCE study group

## Data Availability

data may be made available to scientists upon approval of an application sent to the ENFORCE Scientific Steering Committee and further approval by relevant authorities. Applications for data must be sent to. Detailed information about data access may be found here: https://chip.dk/Research/Studies/ENFORCE/Study-Governance.

## Acknowledgments

The ENFORCE study group members all contributed substantially to the study. A full list of members of the ENFORCE study group is provided as supplementary material.

This study was funded by the Danish Ministry of Health (document 150, parliamentary year 2020/2021, the Danish Parliament).

