## Supplementary material for "Cohort Profile: The Danish National Cohort Study of Effectiveness and Safety of SARS-CoV-2 vaccines (ENFORCE)": The ENFORCE study group

**Sponsor/Data and Statistical Centre:** CHIP, Rigshospitalet under the direction of Professor J Lundgren.

**Principal Investigator:** Professor LJ Østergaard, Department of Infectious Diseases, Aarhus University and Aarhus University Hospital.

**Sites** (alphabetical and regional coordinators in parenthesis)

**Region Hovedstaden:** (T Benfield), L Krohn-Dehli, DK Petersen, Copenhagen University Hospital - Amager and Hvidovre, Hvidovre. K Fogh, EH Mikkelsen, (K Iversen), Copenhagen University Hospital – Gentofte and Herlev, Herlev.

**Region Midtjylland:** P Bek, V Klastrup, F Larsen, SH Rasmussen, MH Schleimann, S Schieber, (NB Stærke), A Søndergaard, B Tarp, M Tousgaard, Y Yehdego, Aarhus University Hospital, Aarhus.

**Region Nordjylland:** J Bodilsen, (H Nielsen), KT Petersen, M Ruwald, RK Thisted, Aalborg University Hospital, Aalborg.

**Region Sjælland:** SF Caspersen, M Iversen, LS Knudsen, JL Meyerhoff, LG Sander, (L Wiese), Zealand University Hospital Roskilde, Roskilde.

**Region Syddanmark:** C Abildgaard, IK Holden, NE Johansen (IS Johansen), L Larsen, SO Lindvig, LW Madsen, A Øvrehus, Odense University Hospital, Odense.

**Scientific Steering Committee:** NA Kruse, H Lomholdt, Lægemiddelstyrelsen, TG Krause, P Valentiner-Branth, Statens Seruminstitut, B Søborg, Sundhedsstyrelsen, TK Fischer, Copenhagen University, C Erikstrup, Aarhus University, SR Ostrowski, Rigshospitalet, H Nielsen, Aalborg University Hospital, IS Johansen, Odense University Hospital, LJ Østergaard (chair), M Tolstrup, NB Stærke, OS Søgaard Aarhus University Hospital, L Wiese, Zealand University Hospital Roskilde, T Benfield, Copenhagen University Hospital – Amager and Hvidovre, J Lundgren, D Raben, CHIP, Rigshospitalet

**Operational group:** H Nielsen, Aalborg University Hospital, IS Johansen, Odense University Hospital, LJ Østergaard, M Tolstrup, NB Stærke, OS Søgaard, Aarhus University Hospital, L Wiese, Zealand University Hospital Roskilde, T Benfield, Copenhagen University Hospital – Amager and Hvidovre, J Lundgren (chair), D Raben, CHIP, Rigshospitalet, E Jylling, Danske Regioner, D Hougaard, Statens Seruminstitut

**Coordinating Centre:** Aarhus University and Aarhus University Hospital, SD Andersen, K Lykkegaard, NB Stærke, OS Søgaard, M Tolstrup, LJ Østergaard

**ENFORCE Lab.:** Aarhus University Hospital: SR Andreasen, E Baerends, LL Dietz, AK Hvidt, AK Juhl, R Olesen, M Tolstrup.

**Data and Statistical Centre:** CHIP, Rigshospitalet: KK Andersen, W Bannister, C Bjernved, TW Elsing, FV Esmann, MA Ghafari, E Gravholdt, SF Jakobsen, ML Jakobsen, CM Jensen, TØ Jensen, D Kristensen, LR Kumar, J Lundgren, C Matthews, N Normand, C Olsson, D Raben, J Reekie, A Traytel, T Weide.

**Other contributors:** AM Hvas, H Støvring, Aarhus University Hospital, Statens Seruminstitut

**Funding:** ENFORCE has received a grant from the Danish Ministry of Health (SUM) (legal deeds 150 28/1 2021 and 263 3/6 2021)
